# Screening for early-stage Alzheimer’s disease using optimized feature sets and machine learning

**DOI:** 10.1101/2020.10.28.20212027

**Authors:** Michael J. Kleiman, Elan Barenholtz, James E. Galvin, for the Alzheimer’s Disease Neuroimaging Initiative

**Affiliations:** Department of Neurology, University of Miami Miller School of Medicine, Miami, FL; Center for Complex Systems and Brain Sciences, Florida Atlantic University, Boca Raton, FL

**Author notes:** Correspondence to: Michael J. Kleiman, Department of Neurology, University of Miami Miller School of Medicine, 1600 NW 10th Ave #1140, Miami, FL 33136. Data used in preparation of this article were obtained from the Alzheimer’s Disease Neuroimaging Initiative (ADNI) database (adni.loni.usc.edu). As such, the investigators within the ADNI contributed to the design and implementation of ADNI and/or provided data but did not participate in analysis or writing of this report. A complete listing of ADNI investigators can be found at: http://adni.loni.usc.edu/wp-content/uploads/how_to_apply/ADNI_Acknowledgement_List.pdf.

**Keywords:** Alzheimer disease, mild cognitive impairment, supervised machine learning, data mining, neuropsychological tests

## Abstract

**Background:** Detecting early-stage Alzheimer’s disease in clinical practice is difficult due to a lack of efficient and easily administered cognitive assessments that are sensitive to very mild impairment, a likely contributor to the high rate of undetected dementia.

**Objective:** Here, we aim to identify groups of cognitive assessment features optimized for detecting mild impairment that can be used in routine screening. We also compare the efficacy of classifying impairment using either a two-class (impaired vs non-impaired) or three-class approach.

**Methods:** Supervised feature selection methods generated groups of cognitive measurements targeting impairment defined at CDR 0.5 and above. Random forest classifiers then generated predictions of impairment for each group using highly stochastic cross-validation, with group outputs examined using general linear models.

**Results:** The strategy of combining impairment levels for two-class classification resulted in significantly higher sensitivities and NPVs, two metrics useful in clinical screening, compared to the three-class approach. Just four neuropsychological features (delayed WAIS Logical Memory, trail-making, patient and informant memory questions), able to be administered in approximately 15 active minutes (∼30 minutes with delay), enabled classification sensitivity of 94.53% (88.43% PPV) with the addition of four more features significantly increasing sensitivity to 95.18% (88.77% PPV) when added to the model as a second classifier.

**Conclusion:** The high detection rate paired with the minimal assessment time of the four identified features may act as an effective starting point when screening for cognitive impairment defined at CDR 0.5 and above.

## INTRODUCTION

Alzheimer’s disease (AD) affects an estimated 6 million Americans [1], although estimates suggest nearly two-thirds of AD cases remain undetected until the latter stages of impairment [2,3]. In the preclinical stages of AD there is early accumulation of neuropathology [4] without detectable effect on cognition or functioning. However, as amyloid and tau deposition increases and neuronal injury and neurodegeneration begins, cognitive decline may be first detected clinically as mild cognitive impairment (MCI) due to AD [5] with relative preservation of activities of daily living followed by progression to clinical AD [6]. Approximately 32% of patients with MCI go on to develop AD within 5 years [7]. While at the present time, there are no disease modifying therapies available, the early detection of AD may empower patients and their families to establish plans, start currently available symptomatic medications, discuss interventions and lifestyle modifications that may provide improved quality of life [8–10], and participate in clinical trials.

Improving the overall accuracy of diagnostic methods often directly leads to improving the detection of early AD cases, especially when multiple modalities (cognitive assessments, biomarkers, genomics) are considered [11]. The challenge to clinicians and researchers is that collecting and measuring these biomarker modalities is resource-intensive, time-extensive, and expensive, especially when targeting early impairment [12–15]. Additionally, it is unclear at this point how much accuracy is gained from extensive biomarker collection or what would necessarily be done differently in the primary care setting [16]. Instead, it may be more effective to focus efforts on improving the cognitive screening process for patients with potential impairments in the primary care setting in order to facilitate the recommendation of further testing or referrals to a specialist for indeterminant or borderline cases. In contrast to diagnosis, where assessment accuracy is paramount, the types of measurements in screening and early detection should be restricted based on resource costs [17]. The most salient resource for many clinics and hospitals is time: a single hour of a physician’s time is worth a considerable amount to a busy clinic or a hospital that may be understaffed or under-resourced. Other than self-report questionnaires (e.g. the Quick Dementia Rating Scale) [18], neuropsychological and neurobehavioral assessments (e.g. Montreal Cognitive Assessment or MoCA) [19] may be the most resource-efficient methods for detecting cognitive impairment [20] with meta-analyses supporting effective screening utility when incorporated into clinical decision support systems that utilize machine learning techniques [21,22].

To improve dementia screening efficacy, it would be useful to identify as small a set of key cognitive measurements (“features”) as possible while still enabling an accurate detection rate. Previous exploration has largely focused on predicting a clinical diagnosis of AD or MCI [23–25] however this approach is potentially misinformative; while neuropsychological and neurobehavioral assessment scores are often highly useful in predictive models of clinical diagnosis, findings may instead reflect that these assessments were used in the process of determining the clinical diagnosis itself. As clinical diagnoses are typically derived from all available evidence of the disorder and are frequently based on the results of any and all cognitive assessments administered, a predictive model that uses clinical diagnosis as its predicted variable should avoid training the model using features that were previously used to determine the diagnosis, lest the model be unrepresentative. Because of this, it may be preferable to use a staging tool such as the Clinical Dementia Rating (CDR) [26] global score as the predicted variable, as the criteria for scoring the CDR is well validated for differentiating between cognitively normal older adults and those with cognitive impairment and scoring is based on a semi-structured interview by an experienced clinician without specifically relying on other assessments or neuropsychological tests [27–29].

In this study, we aim to identify the most useful cognitive assessment features to detect very mild impairment defined by CDR 0.5 and/or mild impairment defined by CDR 1 using data from the Alzheimer’s Disease Neuroimaging Initiative (ADNI). We approach this using supervised feature selection methods guided by random forest algorithms, including a novel implementation that enables two groups of features to be evaluated with the same model. Feature set usefulness is evaluated using high stochasticity cross-validation on classifiers tuned to each set to minimize the impact of any one particular model or iteration, with classification outputs examined using linear models in order to statistically determine the efficacy of each feature set in screening for early-stage cognitive impairment. Furthermore, as degradation of cognitive functioning due to AD is progressive in nature [6] we investigate the utility of combining CDR 0.5 and CDR 1 into a single “impaired” class by comparing its performance with the more traditional approach of keeping CDR stages separate.

## METHODS

### Data

Data used in the preparation of this article was obtained from the Alzheimer’s Disease Neuroimaging Initiative (ADNI) database (adni.loni.usc.edu). The ADNI was launched in 2003 as a public-private partnership, led by Principal Investigator Michael W. Weiner, MD. The primary goal of ADNI has been to test whether neuroimaging or other biomarkers and neuropsychological/clinical assessment can be combined to measure the progression of MCI and early AD. For this study, we examined only neuropsychological assessments, subject and informant questionnaires, and subject demographics.

Data from ADNI is structured in a sparse format, with most measures broken down and recorded granularly. For example, the Mini Mental State Exam (MMSE) [30] question requiring subjects to spell a word backwards is recorded with each individual letter in the word and whether the subject named that letter in the correct place. To facilitate feature selection, these sparse features were recombined where possible using domain knowledge and tests of multicollinearity. In the case of the MMSE’s backwards spelling, the five features were restructured into two: early and late, as the first two letters and last three letters were found to be highly collinear. We also combined the two versions of the trail-making test (Trails A and Trails B) through summation, found the rounded mean average of each of the Everyday Cognition (ECog) scales [31] (keeping the informant and patient scales separate), and found the difference between the final (delayed) trial and the last immediate repetition in the Rey’s Auditory Verbal Learning Test [32], recorded this as a separate feature, and then discarded all of the individual trials. Tests of multicollinearity also indicated a high degree of collineation for the immediate recall and delayed recall for the Wechsler Adult Intelligence Scale (WAIS) Logical Memory test [33]; because including both could negatively impact feature selection, we chose to use the delayed recall option based on findings that delayed recall is affected earlier in AD than immediate recall [34]. A list of all cognitive assessment features used in this study can be found in the main supplemental code directory [35].

After preparation, we first ran back-fill imputation on cognitive assessments collected within six months of baseline testing, to enable to comparison of assessments collected at only marginally different timepoints, and then dropped subjects with data missing in eleven or more features after imputation (∼10% of total 104). We used median imputation to fill in any still missing datapoints with the median value of that feature, effectively removing it from analysis for that subject. Three features (number span forwards/backwards and vegetable verbal fluency) were also removed at this stage for having no non-missing values for any subject.

### Subjects

The main predicted variable for this study was the CDR, measured using the global scale; 0 for normal, 0.5 for questionable or very mild impairment, 1 for mild impairment, 2 for moderate impairment, and 3 for severe impairment. Following preprocessing, we were left with 1566 subjects – 689 with CDR 0, 784 with CDR 0.5, 92 with CDR 1, and one subject with CDR 2. Because analysis cannot occur with a single subject in a given class, only CDRs of 0, 0.5, and 1 were used in this study, leaving us with 1565 total subjects.

### Feature selection

All models were run with either two or three output classes: CDR 0, CDR 0.5, and CDR 1 for the three-class variant and unimpaired (CDR 0) vs impaired (CDR 0.5 and CDR 1) for the two-class variant. To determine the baseline performance of a model with no dimensionality reduction, all available features (the “All-features” set) were analyzed using a series of random forest classifiers: machine learning models that ensemble the results of hundreds of supervised decision trees [36]. Five All-features models were examined; a three-class multiclass model, a two-class normal/impaired model, and three two-class OneVsRest models that each targeted one of the three CDR stages to be ultimately integrated into a novel multi-classifier network.

Feature selection was then conducted for each of the five models on a consistent 25% of the available data (training set), determined using a train/test split algorithm at a constant random state with only the train split used. Each resulting feature set was individually tuned to promote top model performance. Following the selection of each feature set, hyperparameter optimization was achieved using a two-tiered RandomSearch and GridSearch method on 75% of the training set (18.75% of total dataset). Hyperparameters included the number of estimators, maximum percentage of features considered per split, maximum nodes per leaf, minimum samples required per split, maximum depth per tree, and whether bootstrapping was used. Random states for all optimization runs were also kept consistent to ensure reproducibility.

The optimized random forest model from the All-features set was then used within Boruta [37], a model-guided wrapper method for feature selection which functions by statistically comparing the performances of numerous models with successive variables replaced by “shadow” features: copies of the original features with all values randomly shuffled. These shadow features are then collectively compared to each original feature over hundreds of iterations and the features that consistently significantly outperform the collective maximum of the shadow versions are deemed important and thus selected. Features that only marginally outperform the shadow versions, or that outperform at a less than statistically significant rate, are deemed tentative and manually evaluated, and features that underperform are deemed unimportant and not selected.

The metric for feature importance used in the Boruta feature selection algorithm is statistically rigorous but its reliance on repeated random forest models impacts its interpretability. While taking the results of the Boruta model at face value may be acceptable for many applications, our use-case requires as much interpretability as possible, especially when considering tentatively selected features. To maximize interpretability we utilized SHAP (SHapley Additive exPlanations) [38], specifically the TreeExplainer variant implemented in Python [39], which enables all permutations of the Boruta algorithm’s feature importance metrics to essentially be averaged and globally compared. The resulting feature importance metrics allowed us to make decisions on whether tentative features should be included, which selected features were most important, and if features selected or removed by Boruta were justifiable.

### Model generation

While random forest classifiers are natively able to account for three or more classes, only a single feature set is able to be used. Random forests typically determine feature importance by calculating the classification probability of randomly selected features (gini impurity), however this strategy does not perform as well at feature selection compared to other methods (e.g. Boruta), resulting in suboptimal features often being considered for each new branch. To address this, we developed a method based on the technique of model stacking that integrates multiple random forest classifiers, each with their own optimized feature sets, into a single model which we are calling a Multi-Classifier Network (MCN).

For this study, we arrange the MCN to function with a OneVsRest strategy. The first “layer” of an MCN contains one of each classifier that has been previously optimized to target a single class compared to all others. Optimization includes the determination of optimal hyperparameters and features using previous steps as described above in the Feature Selection section. All classifiers receive the same dataset, with further processing performed to include only the selected features and to reclassify labels as either target class (1) or non-target class (0). The output probabilities of each classifier’s target class are appended to an output array and normalized, treated as probabilistic outputs, then cross-validated.

Both two- and three-class models utilized the MCN, with feature sets for each class derived from their previously generated feature sets and optimized classifier models. For example, the three-class Boruta-SHAP model used feature sets optimized for OneVsRest CDR 0, CDR 0.5, and CDR 1. The two-class and three-class models were also investigated using single random forest classifiers, with hyperparameters tuned, to enable comparison with the MCNs. For three-class models, the underrepresented class (CDR 1) was augmented, producing minority classes with sample counts at 50% of the majority class [40]. Categorical variables were accounted for by ensuring augmented values only referenced specified categories. Augmentation was applied to the training sets only after splitting to minimize leakage.

### Analysis

Following the determination of optimal hyperparameters and feature sets for each model, j-split k-repeat high stochasticity cross-validation (j = 20, k = 20) was run using a random forest classifier and custom designed split/repeat cross-validation function [35]. Split/repeat cross-validation is a high stochasticity approach to cross-validation that uses random sub-sampling to split a dataset one or more times, using random or assigned seed values, and then runs models one or more times again using a variable number of random or assigned seed values to influence varied model generation to produce a large number of datapoints that can then be investigated statistically. Splitting the data this way reduces the negative impact of relatively small data, as instead of splitting off a holdout set for testing we instead first split off a smaller training set to use in hyperparameter optimization and feature selection. For overall model evaluation, the larger holdout set is randomly split j-number of times into training/validation/test splits and the smaller training set that has already been used is then integrated back into each training set to ensure no data is wasted and to maintain the integrity of the data. Each split is then run k-number of times with a defined number of random seeds, ensuring that each random forest model produces slightly different results; essentially creating a random forest of random forest models.

The split/repeat cross-validation outputs sensitivity, specificity, positive predictive value, negative predictive value, and accuracy metrics for each class individually as well as measures of the entire model. For two-class models, overall sensitivity is defined traditionally as recall of the target class and overall specificity as recall of the non-target class, however to enable comparison with the three-class models we examine sensitivity based on the weighted mean recall score of each impaired class and specificity as the recall score of the non-impaired class. Positive and negative predictive values are also calculated similarly. These metrics are then integrated into a series of one-way analyses of variance (ANOVA) to statistically examine the variances between and within each model and feature set. Python 3.6 code for feature selection, hyperparameter optimization, classification, cross-validation, statistical analysis, and visualization can be found in [35].

## RESULTS

### Subject Characteristics

The sample consisted of 1565 ADNI participants. There were more men than women overall (819 men, 746 women), with more women represented in the CDR 0 group (306 men, 383 women) and more men represented in the CDR 0.5 (459 men, 325 women) and CDR 1 (54 men, 38 women) groups (*χ*^2^(2, *N* = 1565) = 30.95, *p* < .001). The mean age for the sample was 72.8 ± 7.1 (range 55 - 92). The CDR 1 (74.8 ± 8.7) group was slightly older than both the CDR 0 (72.7 ± 6.1) and CDR 0.5 (72.7 ± 7.6) groups (*F*(2,1562) = 4.01, *p* = .018). Median years of education obtained was “finished college” (16 years) across all groups. A breakdown of sample characteristics can be found in *Table 1*.

**Table 1.** Demographics and characteristics of study population.

|  | CDR 0 | CDR 0.5 | CDR 1 | Three-class<br>p-value | Two-class<br>p-value |
| --- | --- | --- | --- | --- | --- |
| Number of Subjects | 689 | 784 | 92 |  |  |
| Sex, %Female | 55.59% | 41.45% | 41.30% | < .001 | < .001 |
| Age | 72.67<br>(±6.12) | 72.65<br>(±7.61) | 74.81<br>(±8.75) | .018 | .559 |
| Mean Years of Education | 16.68<br>(±2.47) | 16.10<br>(±2.75) | 15.41<br>(±2.62) | < .001 | < .001 |
| MoCA | 25.68<br>(±2.65) | 21.89<br>(±4.53) | 15.67<br>(±4.74) | < .001 | < .001 |
| CDR Sum of Boxes | 0.04<br>(±0.13) | 1.61<br>(±0.98) | 5.58<br>(±1.16) | < .001 | < .001 |
| Memory | 0.00<br>(±0.00) | 0.58<br>(±0.21) | 1.16<br>(±0.38) | < .001 | < .001 |
| Orientation | 0.00<br>(±0.04) | 0.27<br>(±0.32) | 1.07<br>(±0.39) | < .001 | < .001 |
| Judgement/Problem Solving | 0.28<br>(±0.11) | 0.36<br>(±0.28) | 1.01<br>(±0.34) | < .001 | < .001 |
| Community Affairs | 0.00<br>(±0.05) | 0.16<br>(±0.25) | 0.97<br>(±0.40) | < .001 | < .001 |
| Home/Hobbies | 0.00<br>(±0.03) | 0.20<br>(±0.28) | 0.97<br>(±0.36) | < .001 | < .001 |
| Personal Care | 0.00<br>(±0.00) | 0.04<br>(±0.19) | 0.40<br>(±0.49) | < .001 | < .001 |

Chi-squared analysis comparing the CDR with the ADNI consensus diagnoses (cognitively normal, MCI, or AD) determined that a score of 0 on the CDR identified 99.42% (685 of 689) of subjects with no reported impairment, and a score of 0.5 or greater identified 99.89% (875 of 876) of subjects with some form of cognitive impairment (*χ*^2^(4, *N* = 1565) = 2302.66, *p* < .001). All but five subjects diagnosed with MCI were assigned a CDR of 0.5; four were rated as CDR 0 and one was rated as CDR 1. Seventy-six subjects diagnosed with AD were also rated a CDR 0.5. All but one subject with a CDR of 1 were diagnosed with AD.

### Selected models

Fourteen models, including three models with all available features for comparison (All-features), were selected. Half of the models targeted two classes, impaired vs unimpaired, and the remaining half targeted all three CDRs. Of the three-class models, four utilized an MCN based on previously fit OneVsRest random forest models and three used a standard multiclass random forest, with one of each containing the all-feature feature set. The seven two-class models consisted of two MCN models and five standard binary random forests, with one of the standard random forests containing all available features.

One of the two-class and two of the three-class models were comprised of only automatically selected features using Boruta-SHAP, with the three-class models targeting either each individual CDR stage using a OneVsRest approach (MCN) or all three in a multiclass approach. The two-class Boruta-SHAP analysis selected nine total features, while the three-class Boruta-SHAP analysis selected 17 total features. Within the three-class three-classifier MCN, the Boruta-SHAP analysis targeting CDR 0 selected 17 features, CDR 0.5 selected 31 features, and CDR 1 selected 64 features.

Examination of the Boruta-SHAP models revealed that the top four most useful features for detecting impairment were (a) the delayed WAIS Logical Memory test, (b) the sum total of both trail-making tests (A plus B), (c) the subject’s self-report of memory on the ECog, and (d) the ECog’s caregiver report of the subject’s memory. We label this group of features as “Top-4”. In addition to these, the next four most useful features were (e) the immediate recall and (f) delayed recall questions of the ADAS-Cog (questions 1 and 4), (g) the ECog’s divided attention report by the caregiver, and (h) the Functional Activity Questionnaire (FAQ)’s question on remembering appointments, holidays, family occasions, and medications. These top eight features are labeled “Top-8”. The remaining models included the two-class MCN using both the Top-4 and Top-8 groups (Top-4/8 MCN) and the addition of the ECog and/or FAQ features to either the Top-4 or Top-8 feature sets, based on an abundance of both questionnaires in the automatically selected featuresets. Lists of selected sets with their included features can be found in the main supplementary code directory [35].

### Two-class vs three-class approach

We found that examining both CDR 0.5 and CDR 1 using a combined “impaired” category, compared to “unimpaired” CDR 0, was more useful for dementia screening compared to multiclass models that kept each of the three CDR stages separate. Examining the mean performances of the two-class and three-class approaches revealed a significantly greater sensitivity in the two-class Impaired vs Unimpaired (94.38%) grouping compared to the three-class (91.97%) grouping, (*F*(1,5998) = 1839.22, *p* < .001). There were significant differences in positive predictive value (PPV) with the two-class group (88.63%) compared with the three-class (89.45%) group (*F*(1,5598) = 229.81, *p* < .001). When targeting only those who are unimpaired (specificity), the three-class (86.25%) models perform significantly better than the two-class (84.42%) models (*F*(1,5998) = 487.52, *p* < .001). In contrast, the negative predictive values (NPVs) in the two-class (92.23%) group were significantly higher than the three-class (89.57%) group (*F*(1,5998) = 1549.29, *p* < .001). *Table 2* highlights these differences.

**Table 2.** Statistical comparison of two-class and three-class models.

|  | Sensitivity | PPV | Specificity | NPV | Accuracy |
| --- | --- | --- | --- | --- | --- |
| Two-class | **94.38%**<br>($\pm .001$) | 88.63%<br>($\pm .001$) | 84.42%<br>($\pm .001$) | **92.23%**<br>($\pm .001$) | **90.01%**<br>($\pm .001$) |
| Three-class | 91.97%<br>($\pm .001$) | **89.45%**<br>($\pm .001$) | **86.25%**<br>($\pm .001$) | 89.57%<br>($\pm .001$) | 89.44%<br>($\pm .001$) |

Bold indicates significance, $p < .001$ . Confidence intervals are calculated at 95%.

### Model comparison

Overall, analyses reveal that the two-class MCN combining both the Top-4 and Top-8 feature sets, along with both the two-class nine-feature Boruta-SHAP model (addition of Word Recognition section of ADAS-Cog) and the Top-8 model by itself, performed best in detection of CDR 0.5 individuals. However, the Top-4 model was potentially more efficient for screening purposes due to its minimal number of features while still exhibiting a highly accurate detection rate of positive impairment.

Sensitivity for the two-class group indicated that the Top-4/8 MCN (95.18%), 9-feature Boruta-SHAP (94.75%), and Top-8 (94.69%) feature sets were not significantly different from each other (*F*(6,2793) = 53.88, *p* < .001), and thus collectively achieved higher performance than all other feature sets, shown in *Figure 1.a*. The groups with the next highest sensitivities were found to be the Top-8+ECOG+FAQ MCN (94.54%) and Top-4 (94.48%). PPVs had less variance, with all feature sets exhibiting values between 88.32% and 89.14%, however some significant differences were found (*F*(6,2793) = 6.67, *p* < .001); the All-features feature set (89.14%), Top-4/8 MCN (88.77%) and both the Top-8+ECOG+FAQ’s MCN (88.59%) and standard random forest model (88.64%) had the significantly highest PPVs, followed by the 9-feature Boruta-SHAP (88.48%), Top-4 (88.43%) and Top-8 (88.32%) feature sets, seen in *Figure 1.b*. Examining specificity revealed that the All-features model (85.25%), Top-4/8 MCN (84.52%), and both the standard and MCN versions of the Top-8+ECOG+FAQ (84.69% and 84.35% respectively) were significantly higher than other models in the two-class group (*F*(6,2793) = 7.64, *p* < .001) depicted in *Figure 1.c*. The two feature sets in the two-class group with the highest NPVs were the Top-4/8 MCN (93.29%) and the 9-feature Boruta-SHAP (92.67%), *F*(6,2793) = 52.69, *p* < .001 (*Figure 1.d*.).

**Figure 1.**
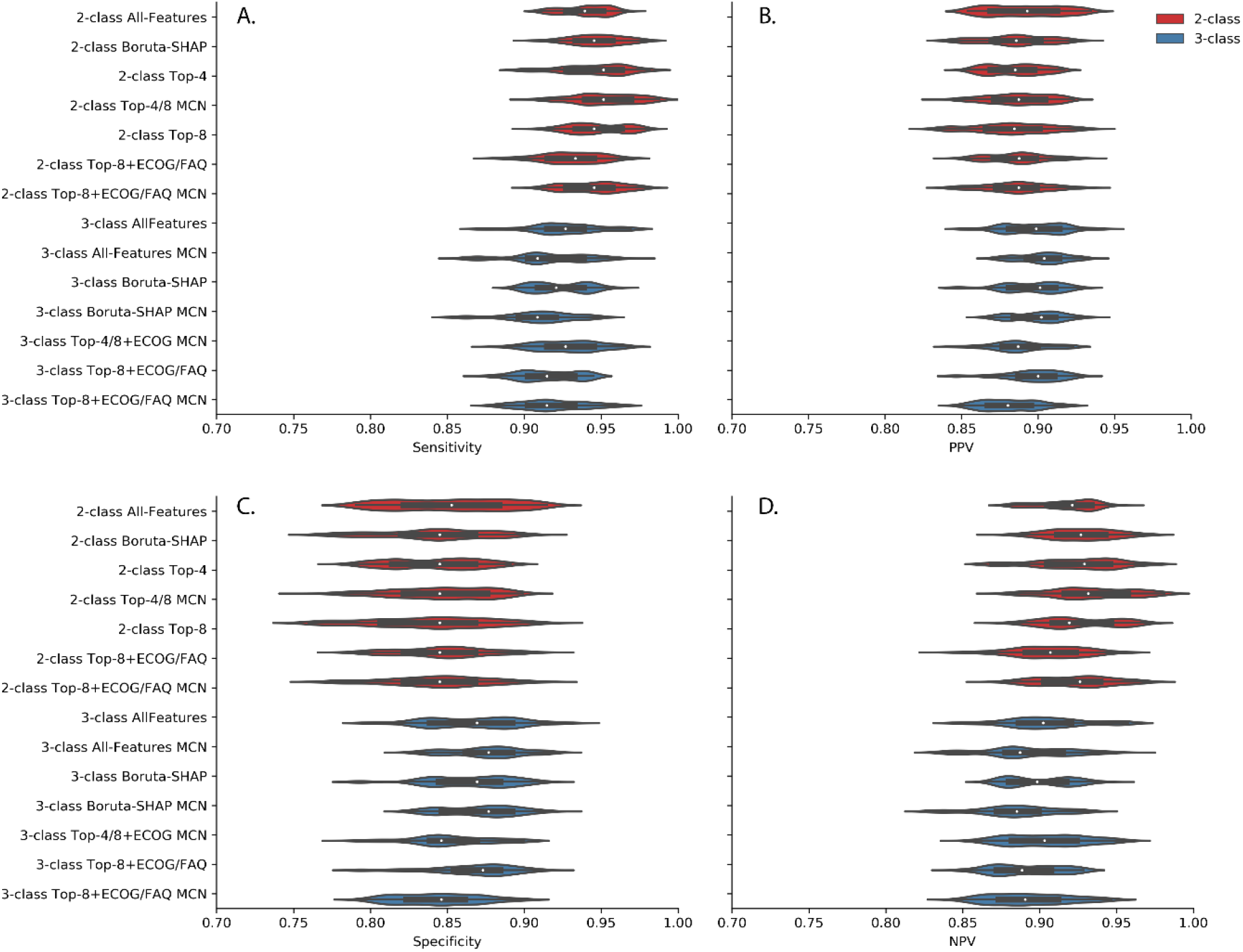
Violin plots displaying the distribution of model outputs for sensitivity (A), positive predictive value (B), specificity (C), and negative predictive value (D). Models that targeted combined impairment (two-class) are depicted in the top half of each subfigure, in red, while models that targeted impairment divided based on severity (three-class) are shown on the bottom half of each subfigure, in blue. Significantly higher sensitivities and negative predictive values can be clearly observed in the two-class models, while the three-class models exhibit significantly higher specificities and positive predictive values.

Within the three-class group, the All-features (92.64%), the 17-feature Boruta-SHAP (92.33%), and the Top-4/8+ECOG MCN (92.84%) feature sets achieved the highest overall sensitivities for the three-class groups (*F*(6,2793) = 39.97, *p* < .001). Similar results were observed for specificity; the All-features MCN (87.53%) and 17/31/64-feature Boruta-SHAP MCN (87.13%) both exhibited significantly higher specificities than other models in the three-class group (*F*(6,2793) = 71.17, *p* < .001). PPVs matched the significances of specificity, with the All-features MCN (90.29%) and 17/31/64-feature Boruta-SHAP MCN (89.94%) having significantly higher values than other feature sets (*F*(6,2793) = 66.36, *p* < .001) and the feature sets with the highest NPVs were the All-features (90.40%), 17-feature Boruta-SHAP (89.98%), and Top-4 MCN (90.46%) (*F*(6,2793) = 34.76, *p* < .001). Sensitivities, specificities, PPVs, and NPVs for all selected models are presented in *Table 3*.

**Table 3.**
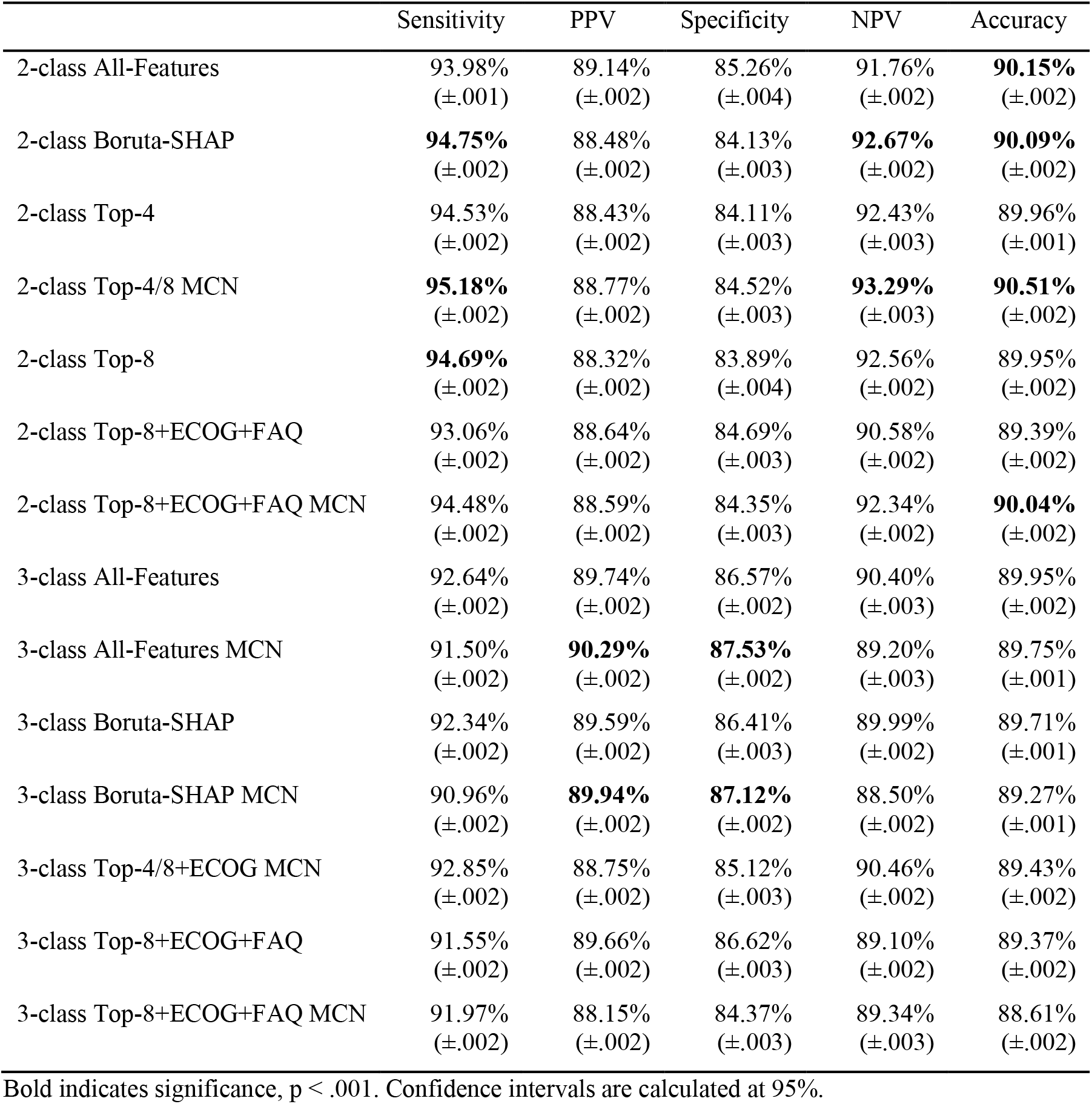
Statistical comparisons of generated feature sets.

## DISCUSSION

Our study identified a minimal feature set that differentiates cognitively normal individuals from those with MCI and mild AD. These minimal feature sets included either four or eight features collected as part of the ADNI protocol; (a) the delayed WAIS Logical Memory test, (b) the sum total of both trail-making tests (A plus B), (c) the subject’s self-report of memory on the ECog, and (d) the ECog’s caregiver report of the subject’s memory comprised the Top-4 features, followed by the inclusion of (e) the immediate recall and (f) delayed recall questions of the ADAS-Cog (questions 1 and 4), (g) the ECog’s divided attention report by the caregiver, and (h) the Functional Activity Questionnaire (FAQ)’s question on remembering appointments, holidays, family occasions, and medications to complete the Top-8 feature set. We further found that combining CDR 0.5 (includes both MCI and very mild AD) and CDR 1 (includes mild AD) into a single “impaired” class before analysis produced significantly higher sensitivities, NPVs, and overall accuracy values compared to models that separated CDR stages. This may be particularly relevant in clinical practice where it can be challenging to differentiate MCI from the mild stages of AD without the advantages of all the biomarkers available in ADNI.

### Impairment classification

For the purposes of detecting clinical disorders, there are differing opinions on whether priority should be given to tests with higher positive detection rates than higher negative detection rates, or vice versa. A test that is optimized towards negative detections (specificity, NPV) allows both differential diagnoses and lack of impairment to be more effectively identified, erring on the side of falsely labeling a positive patient as negative, whereas a test weighted towards positive detections (sensitivity, PPV), erring on the side of falsely labeling a negative patient as positive, may be more useful in cases where a positive identification would lead to further testing. Thus, when screening for potential disorders, a test that leans towards a higher false discovery rate and a lower false omission rate may have more clinical utility, as false positives would likely be correctly diagnosed following further testing whereas false negatives would not be recommended for further testing and miss opportunities for treatment and research participation [41]. We observed lower false omission rates and higher detection rates along with greater overall accuracy in the two-class than the three-class models, suggesting the strategy of combining degrees of impairment may translate to greater clinical utility when screening for AD.

We used the CDR as the predicted measure rather than the consensus clinical diagnosis as using a diagnosis could co-mingle all AD cases in ADNI affecting feature selection for the mildest cases. A potential downside of this is that the CDR is not specific to any one diagnosis, with research finding that a rating of CDR 0.5 may instead identify a range of severities and diagnoses due to different etiologies [42]. Estimating impairment at a level of CDR 0.5 allows for a clear delineation when used for screening purposes. Differential diagnoses can then be determined upon more detailed assessment following a positive result of such screenings.

### Feature sets

With our criteria favoring false positives over false negatives, the two-class Boruta-SHAP and Top-4/8 MCN were the overall best performing models, with similar sensitivity, accuracy, and NPV scores (*Table 3*). While more research is necessary to prospectively confirm clinical utility, the combined use of the delayed WAIS Logical Memory test, the trail-making test, word recall, and informant questionnaires such as the ECog and FAQ appear to be highly useful and efficient for detecting individuals with MCI and mild AD.

It may also be possible to efficiently screen for cognitive impairment by examining only the features in the Top-4 feature set; the delayed WAIS Logical Memory test, the trail-making test, and both the patient’s self-report and informant’s questions of memory functioning on the ECog. This feature set benefits from being highly optimized for estimating CDR-defined impairment with 94.53% sensitivity of detection with a predictive value of 88.43% with the smallest amount of time commitment (estimates of 30 minutes to allow for delayed recall). The ECog and FAQ could be completed prior to the office visit, leaving the office visit for cognitive testing and physical examination. A positive identification based on these key features could then prompt further examination, including collecting the additional measures in the Top-8 feature set or referral for more in-depth neuropsychological, neuroimaging, or biomarker exams.

### Limitations and Future Research

Further research is needed to confirm the clinical utility of these feature sets in independent samples. While many research databases and clinics do not use the exact measures described in this study, it is likely that alternatives can be used provided they measure similar cognitive and behavioral traits. Other patient/informant self-report questionnaires (e.g. the Quick Dementia Rating System [18] or the Informant Questionnaire of Cognitive Function in the Elderly [43]) may prove as useful as the ECog and FAQ, given they ask similar questions, and the WAIS Logical Memory test can be substituted with similar tests of episodic memory with a delayed recall component (e.g. the Craft Story 21 used in the Alzheimer Disease Center’s Neuropsychological Battery in the Uniform Data Set [44]). Likewise, the ADNI database contained a large number of additional neuropsychological tasks, thus future research could incorporate the features identified to be highly useful for impairment detection in this study with novel features or exams to determine potentially more efficient or accurate feature sets for other disorders or stages of disease. Furthermore, while ADNI is considerable in its scope and size, the number of unique patients with relatively complete neuropsychological exam records in the first year of testing is limited. Non-AD dementias were also not included in this study, a step that would be important for the clinical adaptation of these findings.

Results from this study may also be helpful in guiding the development of novel neuropsychological assessments or composite scores. In the case of AD, the inclusion of an episodic memory test with a delayed recall seems to be necessary [12]. The trail-making tests and the informant’s report of patient’s memory were also assigned high importance, promoting the necessity of assessing executive functioning and an informant’s input in detection.

## Conclusions

We found that the use of four neuropsychological features – the delayed WAIS Logical Memory test, trail-making tests A and B, and patient and informant reported memory complaints from the ECog – enable detection of cognitive impairment at 94.53% sensitivity (88.43% PPV) and can serve as an effective starting point when screening for cognitive impairment. Additionally, treating MCI, very mild, and mild AD as a single class instead of separating them may benefit the efficacy of screening tests that utilize machine learning techniques.

## Data Availability

Data used in preparation of this article were obtained from the Alzheimer's Disease Neuroimaging Initiative (ADNI) database (adni.loni.usc.edu). As such, the investigators within the ADNI contributed to the design and implementation of ADNI and/or provided data but did not participate in analysis or writing of this report. A complete listing of ADNI investigators can be found at: http://adni.loni.usc.edu/wp-content/uploads/how_to_apply/ADNI_Acknowledgement_List.pdf

https://github.com/mjkleiman/AD_feature_optimization

## Acknowledgements

Thanks to the members of the University of Miami’s Comprehensive Center for Brain Health, especially Dr. Magdalena Tolea, for providing feedback on the development of this manuscript. Additional thanks to my wife, Lauren Kleiman, for providing feedback on this manuscript even while she works to complete her doctorate.

Data collection and sharing for this project was funded by the Alzheimer’s Disease Neuroimaging Initiative (ADNI) (National Institutes of Health Grant U01 AG024904) and DOD ADNI (Department of Defense award number W81XWH-12-2-0012). ADNI is funded by the National Institute on Aging, the National Institute of Biomedical Imaging and Bioengineering, and through generous contributions from the following: AbbVie, Alzheimer’s Association; Alzheimer’s Drug Discovery Foundation; Araclon Biotech; BioClinica, Inc.; Biogen; Bristol-Myers Squibb Company; CereSpir, Inc.; Cogstate; Eisai Inc.; Elan Pharmaceuticals, Inc.; Eli Lilly and Company; EuroImmun; F. Hoffmann-La Roche Ltd and its affiliated company Genentech, Inc.; Fujirebio; GE Healthcare; IXICO Ltd.; Janssen Alzheimer Immunotherapy Research & Development, LLC.; Johnson & Johnson Pharmaceutical Research & Development LLC.; Lumosity; Lundbeck; Merck & Co., Inc.; Meso Scale Diagnostics, LLC.; NeuroRx Research; Neurotrack Technologies; Novartis Pharmaceuticals Corporation; Pfizer Inc.; Piramal Imaging; Servier; Takeda Pharmaceutical Company; and Transition Therapeutics. The Canadian Institutes of Health Research is providing funds to support ADNI clinical sites in Canada. Private sector contributions are facilitated by the Foundation for the National Institutes of Health (www.fnih.org). The grantee organization is the Northern California Institute for Research and Education, and the study is coordinated by the Alzheimer’s Therapeutic Research Institute at the University of Southern California. ADNI data are disseminated by the Laboratory for Neuro Imaging at the University of Southern California.

